# Domain specific Memory Impairments in Bipolar Mania: Insights from a Tertiary Care Study

**DOI:** 10.1101/2024.09.26.24314411

**Authors:** Mohit Kumar, Sanjay Kumar, Khusboo, Masood Maqbool, Vinit Kumar Singh, Amit Kumar Soni

## Abstract

**Background:** Cognitive deficits in bipolar affective disorder (BPAD), particularly during manic episodes, are well-documented. However, research on domain-specific memory impairments in bipolar mania is limited, especially in the Indian subcontinent. This study aimed to assess memory impairments in patients with bipolar mania using the Postgraduate Institute Memory (PGI-Memory) Scale and to highlight domain-specific deficits compared to healthy controls.

**Methods:** This cross-sectional study was conducted at Tertiary Care Centre in North India. Twenty patients diagnosed with bipolar mania and twenty age, sex and education-matched healthy controls between age 18 to 40 were recruited. Memory functions were assessed using the PGI-Memory scale, focusing on immediate, recent, remote, long-term memory, and associative memory. Mental control and working memory were also evaluated.

**Results:** Both groups were matched in terms of age, sex, and education. the mean (sd) age for bipolar mania group was 27.2 (4.14) years. Patients with bipolar mania demonstrated significant deficits in various memory domains, including immediate, recent, remote, long-term, and associative memory, as well as in visual reproduction and recognition tasks. In contrast, their working memory performance was comparable to that of the control group. The largest deficits were observed in long-term memory (d =2.37) and visual reproduction (d=2.30).

**Conclusions:** Bipolar mania is associated with widespread memory impairments, particularly in long-term and associative memory, which may contribute to difficulties in emotional regulation and daily functioning. These findings emphasize the importance of considering memory impairments in the diagnosis and management of BPAD. Further studies are required to investigate the neurobiological foundations of these impairments and to develop specific interventions.

## Introduction

Bipolar affective disorder (BPAD) is a highly debilitating, chronic, and recurrent mental disorder^1^. It is characterized by a spectrum of extreme mood states, which vary in severity and duration, posing significant challenges in both diagnosis and treatment^2^. According to the World Mental Health (WMH) Survey, the global prevalence of BPAD is estimated at 0.8%^3^.global burden of disease estimated prevalence of BPAD in India is 0.6%^4^. Epidemiological studies suggest that the lifetime prevalence of BPAD may reach as high as 2.4%^3^, leading to significant emotional, social, and economic costs for both families and nations. Despite the fluctuating mood states characteristic of BPAD, patients spend nearly half their lives in depressive or manic episodes, underscoring the need for comprehensive clinical attention to both ends of the mood spectrum.

Mania, a hallmark of BPAD, is characterized by an elevated or irritable mood, excessive energy, increased activity, and impulsive behaviors^5^. These symptoms often result in impairments in cognitive and emotional functioning, affecting job performance, academic work, and social relationships^6^. Individuals in a manic state exhibit heightened distractibility, poor judgment, and difficulty in maintaining organized thoughts. This impaired functioning extends into cognitive domains, often leading to difficulties in everyday life^7–9^.

Cognitive impairment is prevalent in bipolar disorder, particularly during manic episodes^10–13^. Studies have identified deficits in executive functioning, including cognitive flexibility and decision-making, during acute mania ^14,15^. Patients in a manic state show greater impairments in verbal and working memory, executive functioning, reasoning, and problem-solving skills than those who are depressed or in a euthymic phase^16^. These cognitive deficits are linked to the severity of the illness and the number of previous manic episodes^16^. While some cognitive impairments persist during remission, particularly in working memory^14^, many deficits improve with symptom resolution ^17^. However, approximately one-third of bipolar patients may experience persistent cognitive dysfunction even during euthymia^17^.

Research on memory impairment in bipolar disorder, particularly during manic episodes, reveals significant deficits. However, memory is not a singular entity; instead, it consists of multiple distinct systems that vary significantly in their storage durations and capacities^18,19^. Studies consistently show impaired verbal learning and memory in both acute and remission phases^20,21^. During acute mania, working memory deficits are observed, which also persist during remission, while other cognitive domains show mixed results^14^. Notably, verbal retrieval deficits appear to be stable vulnerability indicators, while encoding deficits are specific to manic episodes^22^. A meta-analysis found moderate-to-large impairments across various cognitive domains in euthymic patients, with particularly marked deficits in verbal learning and delayed verbal and nonverbal memory^21^. Similarly, deficits in autobiographical memory and episodic specificity have been observed in individuals with bipolar depression^23^. However, during the manic phase, patients may experience hypermnesia, characterized by an overwhelming number of memories surfacing in a short period ^23^.

However, these findings do not yet provide a comprehensive understanding of the domain specific memory impairments associated with mania. Further research is needed to clarify which aspects of memory are most affected during manic episodes. A clearer memory profile could enhance our understanding of the underlying pathology and inform more targeted psychotherapeutic interventions for patients with bipolar disorder. Hence, this study intended to domain specific memory impairments in bipolar mania.

## Methods

A cross-sectional study was carried out at a tertiary care center in North India using convenience sampling. The study included 20 patients diagnosed with bipolar mania (BM) and 20 healthy controls (HC) matched for age and education. Patients with bipolar mania were recruited from the inpatient services of the hospital, while healthy controls were drawn from the general population. The control group consisted of age- and sex-matched individuals, including bystanders of patients from inpatient or outpatient services, hospital staff, and their family members.

The sample size was determined using G*Power 3 software, aiming for a large effect size of 0.93, an alpha level of 0.05, and a power of 80%. Based on these parameters, a total of 40 participants were required, with 20 individuals in each group.

Individuals diagnosed with Bipolar Affective Disorder according to the International Classification of Diseases—Diagnostic Criteria for Research (ICD-DCR)^24^ criteria, currently undergoing a manic episode lasting at least two months, with or without accompanying psychotic features, and presenting a Young Mania Rating Scale (YMRS) score greater than 15, were recruited for this study. Both male and female participants were eligible if they met the following criteria: aged between 18 and 30 years; right-handed; with a minimum of 8 to 10 years of formal education; literacy in English, or Hindi; and an Intelligence Quotient (IQ) above 70, as assessed by the Wechsler Abbreviated Scale of Intelligence (WASI). Additionally, participants were required to be on stable medication for the preceding month and was not using benzodiazepines.

Patients were excluded if they had more than one prior manic episode, possessed any other Axis I psychiatric disorder—including substance dependence (excluding nicotine)—intellectual disabilities, or identifiable neurological conditions such as epilepsy or dementia. Those with a history of head injury or who had received electroconvulsive therapy (ECT) within the past four months were also not included, as these factors could confound neurocognitive assessments. The study received approval from the Institutional Ethics Committee, and written informed consent was obtained from all participants.

### Assessment/Tools

Patients with bipolar mania were assessed using the Young Mania Rating Scale (YMRS) ^25^, normal controls were evaluated with the General Health Questionnaire–12 (GHQ-12)^26^, and the Edinburgh Handedness Inventory^27^ was administered to both groups. An IQ assessment was conducted using the Wechsler Abbreviated Scale of Intelligence (WASI), and only patients with an IQ greater than 70 were included in the study^28^. The purpose of the IQ test was solely to confirm that scores were not below 70, which is an exclusion criterion for administering the neuropsychological battery. Additionally, the WASI served as a screening tool. The PGI Memory Scale was used to assess domains such as visual retention, immediate and delayed recall, recent memory, and remote memory, in alignment with the domains recommended by the International Society for Bipolar Disorders—Battery for Assessment of Neurocognition (ISBD-BANC) ^29^.

### Young Mania Rating Scale

The Young Mania Rating Scale (YMRS) is one of the most frequently used rating scales to assess mania and was developed by Young et. al. in 1978 ^25^. YMRS is an 11-item instrument designed to assess the severity of manic symptoms in patients already diagnosed with mania. It relies on the patient’s subjective report of their clinical condition over the past 48 hours, supplemented by clinical observations made during the interview. The scale includes four items rated on a 0 to 8-point scale—irritability, speech, thought content, and disruptive/aggressive behavior—and seven items rated on a 0 to 4-point scale. The YMRS has demonstrated strong criterion validity, with a correlation of 0.88 with the Mania Rating Scale, and high inter-rater reliability of 0.93 as reported by the authors.

### Neurocognitive Test Procedures

The Postgraduate Institute Memory (PGI-Memory) Scale is a component of the Postgraduate Institute Battery of Brain Dysfunction (PGI-BBD) ^30^, which includes five batteries: the Performance Intelligence Battery, Verbal Intelligence Battery, Nahor Benson Test, and Bender Gestalt Test and PGI-Memory Scale. The PGI-Memory Scale evaluates various aspects of memory, including remote, recent, and immediate memory, using both verbal and non-verbal materials. It consists of 10 subtests that have been standardized for use in adult populations. The scale has demonstrated strong reliability, with test-retest reliability over a one-week period ranging from 0.69 to 0.85 for the individual subtests and approximately 0.90 for the total scale (both test-retest and split-half reliability). Additionally, the PGI-Memory Scale has shown significant correlations with other established memory assessments, including a 0.71 correlation with the Boston Memory Scale and a 0.85 correlation with the Wechsler Memory Scale. A summary of the names and procedures for each subtest is provided in Table 1.

**Table 1:** Tests and Procedures of PGI Memory Scale.

| S.N. | Test Name | Measures | Procedure |
| --- | --- | --- | --- |
| 1 | Remote Memory | Remote Memory | Questions about significant life events (e.g., when did you get married, pass high school, youngest sibling's name) similar to Levine and colleagues<sup>31</sup> |
| 2 | Recent Memory | Recent Memory | Questions like "What did you eat last night?", "What month is it?", "What day is it today?" |
| 3 | Mental Balance | Mental control | Reverse counting from 20, or performing series subtraction by 3 |
| 4 | Attention and Concentration | Verbal working memory | Forward and backward digit span tasks |
| 5 | Delayed Recall | Long-Term Memory | Present a list of words and ask the participant to recall them after a delay |
| 6 | Immediate Recall | Immediate memory | Sentence repetition with variable lengths and levels of difficulty<sup>32</sup> ranging from 3 syllables to 24 syllables. |
| 7 | Verbal Retention for Similar Pairs | Associative Memory | Pair associative learning for similar word pairs |
| 8 | Verbal Retention for Dissimilar Pairs | Associative Memory | Pair associative learning for dissimilar word pairs |
| 9 | Visual Retention | Visual reproduction | Show a card with a pattern; participant has to draw the same pattern after 15 seconds |
| 10 | Recognition | Recognition Memory | Show a card with 10 objects for 30 seconds, then ask the participant to identify them from a 20-item card |

### Statistical Analysis

To compare the neurocognitive test scores between the two groups, we conducted several statistical analyses. Initially, the Shapiro-Wilk test was applied to assess the normality of data distribution, and Levene’s test was used to evaluate the homogeneity of variances. The results from these tests indicated that the data satisfied the assumptions necessary for parametric testing. Therefore, we utilized independent sample t-tests to compare the mean scores between the bipolar mania group and the control group. Effect sizes were calculated using Cohen’s d, interpreting values as small (d = 0.2), medium (d = 0.5), or large (d = 0.8) effects^33^. For categorical variables, the chi-square test was employed to determine any significant differences between groups. These statistical methods ensured a rigorous comparison of neurocognitive performance across all examined domains.

## Results

### Sample Characteristics

All 40 participants—20 individuals in the bipolar mania group and 20 in the control group— successfully completed the clinical and neuropsychological assessments. The mean age of the entire sample was 27.2 years with a standard deviation of 4.14, indicating a relatively homogenous age distribution among participants. The gender distribution was predominantly male, with 75% males and 25% females in both groups. The demographic and sociocultural characteristics of the bipolar mania group and the control group are summarized in the table 2. Both groups were perfectly matched for sex (75% male and 25% female, p = 1.000), religion (85% Hindu, p = 1.000), and age (mean 27.2 years, p = 1.000). There were no significant differences in family type (p = 0.429), marital status (p = 0.519), or education level (mean years of education: 11.2 for the bipolar group and 10.67 for the control group, p = 0.951).

**Table 2.** Comparison of Demographic and Sociocultural Variables Between Bipolar Mania and Control Groups.

| Variables | | Bipolar mania Group | Control Group | t/ $\chi^2$ | P |
| --- | --- | --- | --- | --- | --- |
|  |  | Mean (SD) or n (%) | Mean (SD) or n (%) |  |  |
| Sex | Male | 15 (75) | 15 (75) | .000 | 1.000 |
|  | Female | 5 (25) | 5 (25) |  |  |
| Religion | Hindu | 17 (85) | 17 (85) | .000 | 1.000 |
|  | Others | 3 (15) | 3 (15) |  |  |
| Habitat | Rural | 12 (60) | 8(40) | 1.600 | 0.206 |
|  | Urban | 8(40) | 12(60) |  |  |
| Family Type | Nuclear | 5 (25) | 3(15) | 0.625 | 0.429 |
|  | Joint | 15 (75) | 17(85) |  |  |
| Occupation | Employed | 0 (0) | 4(20) | 4.444 | .035* |
|  | Un-employed | 20 (100) | 16(80) |  |  |
| Marital status | Married | 7 (35) | 9(45) | 0.417 | 0.519 |
|  | Un-married | 13 (65) | 11(55) |  |  |
| Age |  | 27.2 (4.14) | 27.2 (4.14) | 0.000 | 1.000 |
| Education |  | 11.2 (2.09) | 10.67 (2.02) | 1.010 | 0.951 |

\*p< .05

However, a significant difference was observed in occupational status between the two groups. All participants in the bipolar mania group were unemployed, while 20% of the control group were employed. This finding suggests a potential association between bipolar mania and unemployment. In contrast, no significant difference was found regarding habitat: 60% of individuals in the bipolar mania group lived in rural areas compared to 40% in the control group. This indicates that the distribution of rural and urban residences was relatively similar between the groups.

### Clinical Characteristics of the Bipolar-Mania Group

The mean age of the participants was 27.2 (4.14) years, with most experiencing the onset of bipolar disorder in early adulthood, at a mean age of 22.46 (2.97) years. Three patients had a history of depression prior to the current manic episode. The patients had manic episodes lasting an average of 4.04 months (SD = 3.48), and the mean duration of illness was 6.12 months (SD = 2.54). The average score on the Young Mania Rating Scale (YMRS) was 23.21 (SD = 4.38), indicating the severity of their manic symptoms. Of the 20 patients, only two were on mood stabilizers, while the remaining 18 were prescribed atypical antipsychotics.

### Domains of Memory

Table 3 summarizes the neurocognitive performance differences between the bipolar mania group and the control group across various memory domains. In the domain of immediate, recent, and remote memory, the bipolar group exhibited significantly lower performance than the control group. For remote memory, the bipolar group scored an average of 4.45 (SD = 1.39) compared to 6.00 (SD = 0.00) in the control group, showing a large effect size (Cohen’s d = 1.58, p < .001). Similarly, recent memory performance in the bipolar group (M = 4.35, SD = 0.99) was significantly lower than the control group (M = 5.00, SD = 0.00), with a medium effect size (Cohen’s d = 0.93, p = .006). Immediate memory also showed a marked deficit in the bipolar group (M = 6.05, SD = 2.39) relative to the control group (M = 9.30, SD = 2.00), with a large effect size (Cohen’s d = 1.47, p < .001).

**Table 3.** Comparison of Memory Domain Performance Between Bipolar Mania and Control Groups.

| Domains | Bipolar-mania<br>Group | Control<br>Group | t | P | Effect<br>Size |
| --- | --- | --- | --- | --- | --- |

|  |  |  |  |  | (Cohen's d) |
| --- | --- | --- | --- | --- | --- |
| Remote Memory | 4.45 ± 1.39 | 6.00 ± 0.00 | -4.98 | < .001*** | 1.58 |
| Recent Memory | 4.35 ± 0.99 | 5.00 ± 0.00 | -2.95 | .006** | 0.93 |
| Mental control | 5.85 ± 1.53 | 7.80 ± 2.01 | -3.45 | .001*** | 1.8 |
| Verbal working memory | 8.00 ± 1.21 | 8.25 ± 0.96 | -0.73 | 0.476 | 0.23 |
| Long-Term Memory | 6.90 ± 1.48 | 9.70 ± 0.80 | -7.43 | < .001*** | 2.37 |
| Immediate memory | 6.05 ± 2.39 | 9.30 ± 2.00 | -4.66 | < .001*** | 1.47 |
| Associative Memory | 3.65 ± 1.26 | 4.95 ± 0.22 | -4.52 | < .001*** | 1.4 |
| Associative Memory | 6.30 ± 4.15 | 12.60 ± 2.52 | -5.8 | < .001*** | 1.84 |
| Visual reproduction | 8.30 ± 2.25 | 12.30 ± 0.97 | 7.3 | < .001*** | 2.3 |
| Recognition Memory | 7.55 ± 2.06 | 9.90 ± 0.44 | -4.98 | < .001*** | 1.54 |

\*\*\*p<.001, \*\*p<.01

In the domains of mental control and working memory, the bipolar group demonstrated significant impairments. For mental control, the bipolar group scored lower (M = 5.85, SD = 1.53) compared to the control group (M = 7.80, SD = 2.01), with a significant difference (Cohen’s d = 1.80, p = .001). However, no significant difference was observed in verbal working memory, where the bipolar group (M = 8.00, SD = 1.21) and the control group (M = 8.25, SD = 0.96) showed comparable performance (Cohen’s d = 0.23, p = 0.476).

When assessing learning and associative memory, significant deficits were observed in the bipolar group. In long-term memory, the bipolar group scored an average of 6.90 (SD = 1.48), significantly lower than the control group (M = 9.70, SD = 0.80), with a very large effect size (Cohen’s d = 2.37, p < .001). Associative memory performance was also impaired in the bipolar group, with scores of 3.65 (SD = 1.26) for similar pairs and 6.30 (SD = 4.15) for dissimilar pairs, compared to 4.95 (SD = 0.22) and 12.60 (SD = 2.52) in the control group, respectively. Both comparisons showed large effect sizes (Cohen’s d = 1.4 and 1.84, both p < .001).

In terms of visual reproduction and recognition memory, the bipolar group exhibited considerable deficits. For visual reproduction, the bipolar group scored significantly lower (M = 8.30, SD = 2.25) than the control group (M = 12.30, SD = 0.97), with a very large effect size (Cohen’s d = 2.3, p < .001). Similarly, in recognition memory, the bipolar group (M = 7.55, SD = 2.06) performed significantly worse than the control group (M = 9.90, SD = 0.44), with a large effect size (Cohen’s d = 1.54, p < .001).

All memory domains in the bipolar mania group were impaired compared to the control group, except for working memory, which showed no significant difference between the groups. As depicted in Figure 1, long-term memory exhibited the greatest impairment, followed by visual retention. In contrast, recent memory was among the least impaired domains. Despite the widespread cognitive deficits, working memory remained relatively unaffected in comparison to controls.

**Figure 1.**
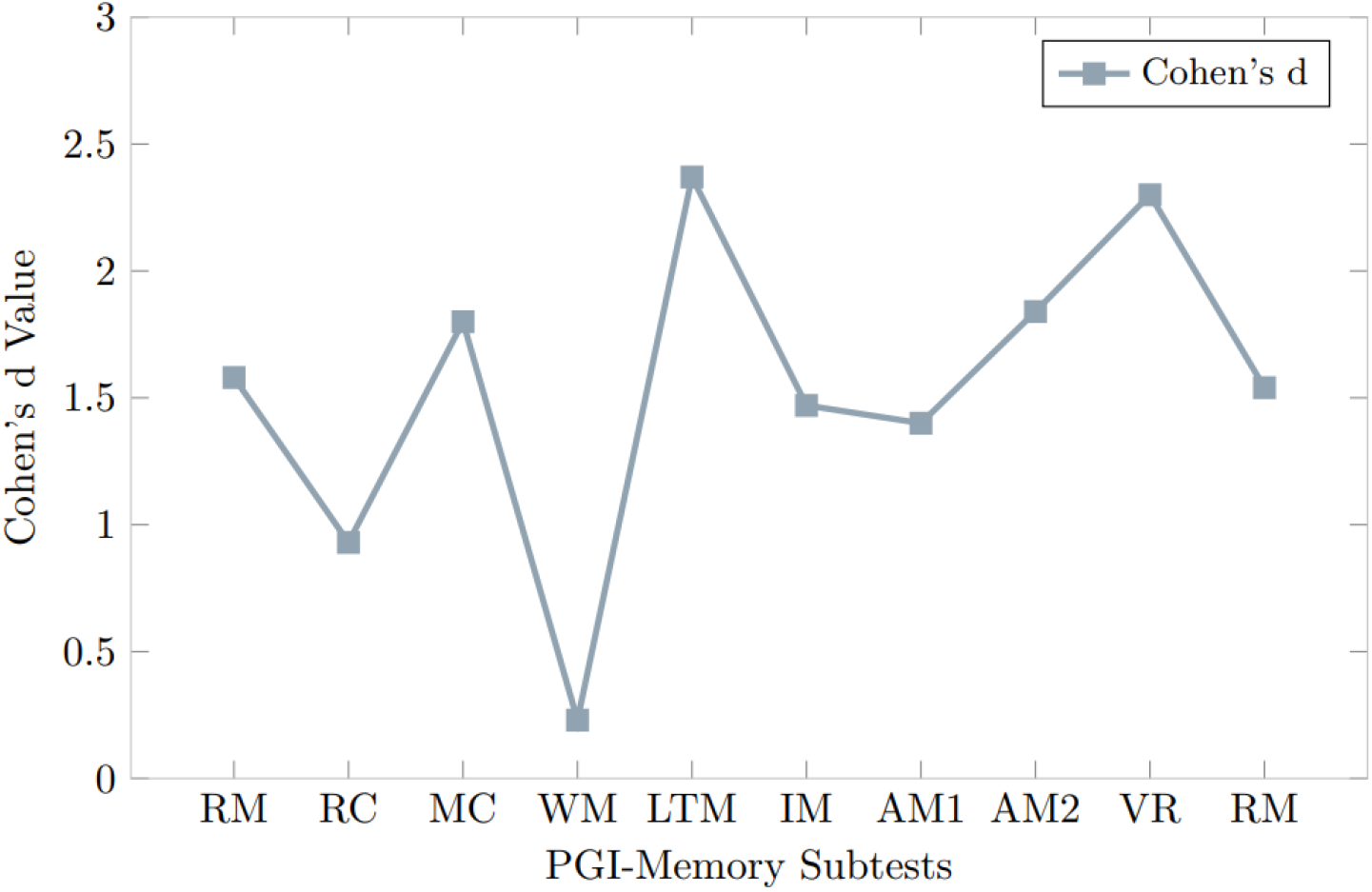
Cohen’s d Values for PGI-Memory Subtests Comparing Bipolar and Control Groups. Note: The following abbreviations represent the memory domains assessed using the PGI Memory Scale: RM = Remote Memory; RC = Recent Memory; MC = Mental Control; WM = Working memory; LTM = Long-Term Memory; IM = Immediate Memory; AM1 = Associative Memory for Similar Pairs; AM2 = Associative Memory for Dissimilar Pairs; VR = Visual Reproduction; RN = Recognition.

## Discussion

The present study investigated domain specific memory impairment in Bipolar affective disorder current episode mania utilizing the Postgraduate Institute Memory (PGI-Memory) scale. While previous research has explored cognitive functioning in various phases of BPAD, including depression, euthymia, and mania, whereas domain-specific memory impairments in bipolar mania have not been extensively studied. Our study revealed significant deficits in almost all domains of memory including immediate, recent, remote, and associative memory, as well as long-term memory, visual reproduction and recognition memory in individuals with bipolar mania, as compared to healthy controls. Importantly, working memory appeared to be unaffected in the bipolar-mania group, suggesting domain-specific memory impairments associated with the manic phase of bipolar disorder.

The bipolar mania group and the control group were largely comparable across most socio-demographic variables, including sex, religion, habitat, family type, marital status, age, and education level, suggesting these factors are unlikely to confound the cognitive outcomes observed. However, none of the patients in the bipolar mania group were employed, compared to 20% employment in the control group. This disparity may result from poor cognitive abilities associated with bipolar mania, impacting employment due to impaired judgment, heightened impulsivity, and difficulties in social interactions^10,34^. Supporting our findings, previous studies have demonstrated that cognitive impairments in bipolar disorder significantly predict occupational recovery and employment status, highlighting the importance of addressing cognitive deficits to enhance vocational functioning and quality of life in individuals with bipolar mania^35,36^.

Mood state at the time of encoding, whether manic, depressed, or euthymic, plays a significant role in influencing the ability to recall events later. We hypothesized that patients in a manic state would exhibit poor recollection of remote events, and our findings confirmed that individuals with mania showed impairments in immediate, recent, and remote memory. Previous research on autobiographical memory^23,37^, a subset of remote memory, has highlighted similar deficits particularly in episodic memory. One of the studies found that patients with mania displayed greater episodic memory impairments compared to euthymic or depressed individuals, whereas semantic memory remained relatively unaffected^23^. Consistent with these findings, we observed that remote memory was more impaired than recent memory in patients with bipolar mania, suggesting a particular vulnerability in episodic memory during manic episodes. The remote memory questions in the PGI Memory Scale primarily assess episodic memory, as they require recall of personally experienced events from the past. This specificity further supports the pattern of episodic memory impairment observed in our study. Additionally, the impairments in immediate and recent memory observed in our study suggest that the deficits in mania are likely due to encoding difficulties rather than issues with storage or retrieval, a conclusion that is in line with other studies highlighting encoding impairments in patients with mania^37^.

Several previous studies have identified abnormalities in the structure and function of the hippocampus in patients with bipolar disorder, which may explain the memory impairments observed during mania^38–40^. According to the multiple trace theory, the hippocampus is essential not only for encoding events but also for retrieving them, particularly in the case of episodic memory^41^. This theory posits that the hippocampus acts as a pointer, directing attention to relevant information during memory recall. In contrast, semantic memory is less reliant on the hippocampus^42^. Our findings, which show greater impairment in episodic rather than semantic memory, align with this theory. Additionally, fMRI and other neuroimaging studies have corroborated these results, showing disrupted hippocampal activity in individuals with bipolar disorder during memory tasks. This hippocampal dysfunction likely contributes to the encoding deficits we observed in immediate and recent memory, as well as the retrieval difficulties seen in remote memory^43–46^.

Further, mental control and working memory are well-studied in BPAD, with mental control involving the regulation of cognitive processes like attention and task-shifting, while working memory is the temporary storage of verbal or visual information^19^. Our study reveals that patients with bipolar mania had impaired mental control but intact working memory, as measured by the digit span task. This contrasts with previous research that frequently reports working memory impairments in BPAD^21,47–50^. However, studies that account for factors such as inhibitory control^51^ suggest that working memory may appear intact when attention and inhibitory control are effectively managed. In line with Larson et al. (2005), the impaired mental control we observed could be due to deficits in inhibitory control, leading to poor self-regulation during manic episodes. Baddeley’s Hedonic Detector^52^ further explains this by highlighting the dysfunctional interaction between emotional processing and working memory in bipolar disorder, which could explain the specific deficits in mental control without affecting basic working memory capacity.

In line with previous research^20,47–49,53,54^, our study found broad-based memory deficits in patients with mania, with the most prominent impairments observed in long-term verbal memory and recognition memory. Specifically, patients showed significant difficulties in both verbal recall, visual immediate reproduction and visual recognition, suggesting that the encoding and retrieval processes are compromised during manic episodes. Similar findings have been reported in studies using the California Verbal Learning Test (CVLT), and Rey-Osterrieth Complex Figure Test (ROCF) where patients with BPAD demonstrated poor performance on both verbal memory and visual memory ^53^. These deficits in long-term memory and recognition may reflect underlying dysfunctions in the hippocampus and prefrontal cortex ^55^, which are crucial for memory consolidation and retrieval, and may contribute to the overall cognitive impairments observed in mania

Our study found significant impairments in pair association learning in patients with mania, indicating a marked deficit in their ability to form and retrieve associations between stimuli. These findings suggest a disruption in the consolidation and retrieval of associative memories—a process heavily dependent on the hippocampus and prefrontal cortex, both of which are essential for memory encoding and executive functions^56,57^. According to Friston’s free energy principle^58,59^, the brain minimizes uncertainty by forming associations between sensory inputs and stored information. In mania, the dysregulated neural activity in these key brain regions may disrupt this process, leading to difficulties in associative learning.

The present study highlights a broad range of memory impairments in bipolar mania, which are crucial for emotional regulation, decision-making, and daily functioning. One of the strengths of our study is the careful matching of the bipolar mania group and the control group for age, sex, and education. This matching minimizes potential confounding variables and enhances the validity of our findings. These findings can enhance clinicians’ ability to conduct mental status examinations and improve diagnostic accuracy by focusing on how memory details are generated across different mood states. For example, our results indicate that memory impairments during mania are primarily related to difficulties in encoding information. In contrast, depression tends to affect recall ability, resulting in memories with fewer perceptual details and fragmented timelines^23^. This distinction is important in clinical assessments, particularly when asking patients about the sequence of events, as it can guide more tailored diagnostic and therapeutic approaches. Episodic memory deficits may contribute to repeated maladaptive behaviors, while mental control impairments suggest difficulties in regulating thoughts and attention, leading to impulsive reactions. Recognizing these specific deficits could provide clear clinical provide and guide the development of targeted interventions to improve memory consolidation and executive control in individuals with bipolar disorder. Understanding the precise memory impairments in bipolar mania can also lead to more effective clinical interventions for managing cognitive dysfunctions.

One limitation of this study is the relatively small sample size and the exclusive focus on patients in the manic phase of bipolar disorder, which may limit the generalizability of the findings. Future research should aim to replicate these results with larger and more diverse populations, including individuals in depressive and euthymic states, to enhance the applicability of the conclusions. Additionally, the cross-sectional design limits our ability to determine whether cognitive impairments persist across mood states or vary with symptom severity. Longitudinal studies are needed to track cognitive changes over time in bipolar disorder. Although we used an Indian neuropsychological battery, future studies should consider employing more comprehensive cognitive assessment tools. Lastly, the study did not account for possible confounding variables such as medication status or the presence of comorbid conditions, which could have influenced the cognitive performance of the participants.

## Conclusion

In conclusion, this study identified significant domain-specific memory impairments in patients with bipolar mania, particularly in visual and verbal memory, episodic memory, mental control and associative learning, while working memory remained unaffected. These findings suggest that cognitive dysfunction in BPAD is selective and linked to underlying neural abnormalities, particularly in the hippocampus and frontal-subcortical circuits. Understanding these impairments provides valuable insight into the cognitive challenges faced by individuals with bipolar disorder and underscores the importance of developing targeted interventions to improve memory and executive functions in this population. Further studies are needed to investigate the neurobiological mechanisms that contribute to these deficits and to assess the effectiveness of cognitive interventions in addressing these impairments.

## Data Availability

All data produced in the present study are available upon reasonable request to the authors

